# COVID-19 in people with neurofibromatosis 1, neurofibromatosis 2, or schwannomatosis

**DOI:** 10.1101/2022.03.31.22273208

**Authors:** Jineta Banerjee, Jan M. Friedman, Laura J. Klesse, Kaleb Yohay, Children’s Tumor Foundation Clinical Care Advisory Board, Justin T Jordan, Scott Plotkin, Robert J Allaway, Jaishri Blakeley

**Affiliations:** Sage Bionetworks, Seattle, WA; Department of Medical Genetics, Faculty of Medicine, University of British Columbia, Vancouver, BC; Department of Pediatrics and Neurosurgery, Simmons Cancer Center, University of Texas Southwestern Medical Center, Dallas, TX; Comprehensive Neurofibromatosis Center at NYU Langone Health, New York, NY; Department of Neurology, Massachusetts General Hospital, Boston, MA; Stephen E. and Catherine Pappas Center for Neuro-Oncology, Massachusetts General Hospital, Boston, MA; Departments of Neurology, Oncology, and Neurosurgery, Johns Hopkins University School of Medicine, Baltimore, MD

## Abstract

**Purpose:** People with pre-existing conditions may be more susceptible to severe Coronavirus disease 2019 (COVID-19) when infected by severe acute respiratory syndrome coronavirus-2 (SARS-CoV-2). The relative risk and severity of SARS-CoV-2 infection in people with rare diseases like neurofibromatosis (NF) type 1 (NF1), neurofibromatosis type 2 (NF2), or schwannomatosis (SWN) is unknown.

**Methods:** We investigated the proportions of SARS-CoV-2 positive or COVID-19 patients in people with NF1, NF2, or SWN in the National COVID Collaborative Cohort (N3C) electronic health record dataset.

**Results:** The cohort sizes in N3C were 2,501 (NF1), 665 (NF2), and 762 (SWN). We compared these to N3C cohorts of other rare disease patients (98 - 9844 individuals) and the general non-NF population of 5.6 million. The site- and age-adjusted proportion of people with NF1, NF2, or SWN who tested positive for SARS-CoV-2 or were COVID-19 patients (collectively termed *positive cases*) was not significantly higher than in individuals without NF or other selected rare diseases. There were no severe outcomes reported in the NF2 or SWN cohorts. The proportion of patients experiencing severe outcomes was no greater for people with NF1 than in cohorts with other rare diseases or the general population.

**Conclusion:** Having NF1, NF2, or SWN does not appear to increase the risk of being SARS-CoV-2 positive or of being a COVID-19 patient, or of developing severe complications from SARS-CoV-2.

## INTRODUCTION

Neurofibromatosis type 1 (NF1), neurofibromatosis type 2 (NF2) and schwannomatosis (SWN) are autosomal dominant genetic conditions predisposing patients to tumors involving the central and peripheral nervous system. NF1 is much more common (estimated prevalence of 1/3600) than NF2 (1/56,000) or schwannomatosis (1/126,000).^1,2^ Given that NF1, NF2, and SWN often cause chronic health impairments, the care community has been concerned about the possibility of increased risk of infection or severe outcomes of COVID-19 in people with one of these genetic conditions. For instance, NF1 is associated with several types of malignant tumors (e.g., malignant peripheral nerve sheath tumors, juvenile myelomonocytic leukemia and glioma), non-malignant tumors, and a range of other manifestations (e.g., vasculopathy and cognitive deficits). People with NF1 have a reduced life expectancy attributed predominantly to premature death caused by cancer or vasculopathy.^3,4^ Some of these manifestations might increase risks associated with SARS-CoV-2 infection. Additionally, while people with NF1 have been impacted by access to routine care and delayed activity in clinical trials during the COVID-19 pandemic,^5,6^ it is unknown if people with NF1, NF2, or SWN are more susceptible to severe acute respiratory syndrome-coronavirus-2 (SARS-CoV-2) infection or if they are more likely to have severe symptoms of the disease than other populations.

To address these questions, we explored the health records of people with NF1, NF2, or SWN in the National COVID Cohort Collaborative (N3C) Data Enclave^7^ to estimate the proportion of patients with these diagnoses affected by SARS-CoV-2 or COVID-19. The N3C Enclave is a dataset and analysis platform that permits researchers to access, query, and analyze COVID-19-related electronic health record data (including standardized clinical diagnoses, laboratory results, medication records, procedures, and visit records) from 55 participating healthcare sites and an estimated 6.4 million individuals in the United States (to July 2021)^7,8^ to better understand the impact of COVID-19 on specific populations.

This study explores the proportions of SARS-CoV-2 positive or COVID-19 positive patients in people with NF1, NF2, and SWN. Further, we examine the proportions of positive cases with NF1 who experienced high severity of COVID-19 disease based on the retrospective observational data available in N3C.

## MATERIALS AND METHODS

## Data access

Data access and analysis in this study were compliant with a research protocol (Sage Bionetworks #2021101002) approved by the Western Institutional Review Board - Copernicus Group (WCG) IRB and granted IRB-exempt status. A request (#RP-DD0EDC) for access to the de-identified N3C dataset (phenotypic acquisition v3.3) was submitted and approved by the N3C Enclave Data Access Committee. All cohorts were generated using custom SQL queries within the N3C Data Enclave and subsequently analyzed using Contour, Fusion, R and Python in the N3C computing environment. Some of the aggregate data were downloaded after review and approval by the N3C data access committee and further analyzed and visualized in a private, secure cloud computing instance provisioned by Sage Bionetworks. The data analyzed in this study were last updated on July 29, 2021.

### SARS-CoV-2 positive or COVID-19 patient criteria

Patients were documented as SARS-CoV-2 positive or COVID-19 patients (together called *positive cases*) in the N3C Data Enclave if they had a hospital visit after 1/1/2020 and had one or more of the following: 1) a positive result from one or more of a set of predefined SARS-CoV-2 laboratory tests, 2) a “strong positive” COVID-19 diagnostic code from the ICD-10 or SNOMED tables described in Version 3.3 of the N3C Phenotype Documentation^9^, or 3) two “weak positive” COVID-19 diagnostic codes from the ICD-10 or SNOMED tables in the phenotype documentation during the same encounter or on the same date prior to 5/1/2020. In cases where a patient had both a positive laboratory test result and a positive COVID-19 diagnosis code, priority was given to the criteria that had an earlier date. In cases where a patient had both positive test results and COVID-19 diagnosis code documented on the same date, priority was given to the positive laboratory test result. Since one criterion was selected and documented for each patient to determine their SARS-CoV-2 or COVID-19 status, there was no duplication of individuals if they satisfied more than one criterion.

Each positive case entered in N3C was matched to two SARS-CoV-2-negative patients *(controls)* at the same site by age, sex, and race. Patients were considered SARS-CoV-2 negative *(controls)* if they met any of the following criteria: 1) set of predefined SARS-CoV-2 laboratory tests with a non-positive result, 2) did not qualify as a COVID-19 patient, or 3) had at least 10 days between the minimum and maximum encounter date in the electronic health record (EHR), to eliminate patients who were only seen for a COVID test. The N3C cohort definition and positive and negative case criteria are publicly available as described in version 3.3 of the N3C COVID-19 Phenotype Documentation (https://github.com/National-COVID-Cohort-Collaborative/Phenotype_Data_Acquisition/wiki/Latest-Phenotype).

### Patient cohort selection

The patients included in the N3C dataset consist of positive cases and control patients from same contributing sites in the ratio 1:2, matched by age, gender (Female, Male, Other), race (White, Black or African-American, Native Hawaiian or Pacific Islander, Asian, Other, Missing/Unknown) and ethnicity (Hispanic, Non-Hispanic, Missing/Unknown) as per version 3.3 of the N3C COVID-19 Phenotype Documentation.^8^

In this study, the dataset was stratified by disease, selecting several disease cohorts to use as comparison groups. An NF1-specific concept set was constructed using neurofibromatosis type 1-relevant diagnosis codes (Supplemental Table 1) from SNOMED, ICD9/10, LOINC, and Nebraska Lexicon (N3C Codeset ID: 792972142). Any unique person in the N3C dataset (identified by their unique N3C person ID) who had diagnosis codes belonging to any of the NF1-relevant concepts defined by the OMOP (Observational Medical Outcomes Partnership) common data model (24 concepts; Supplemental Table 1) was included in the NF1 cohort. This was repeated for neurofibromatosis type 2 (NF2) and schwannomatosis (SWN). Our comparison cohorts also included patients with other rare diseases like fragile X syndrome (FX), tuberous sclerosis (TS), Merkel cell carcinoma (MCC), or acute myeloid leukemia (AML); as well as non-rare diseases like diabetes mellitus type 1 (DM1) or controlled hypertension (HYP), the N3C population without NF1 (Non-NF1), without NF2 (Non-NF2), or without SWN (Non-SWN). Concept sets for these diseases are available in Supplemental Tables 1-9. Any patient with missing data for pre-existing diagnosis, SARS-CoV-2 test, COVID-19 diagnosis, or age was excluded from the analysis. Due to data anonymization prior to contribution to the N3C database, if a patient visited more than one of the 55 healthcare sites contributing to N3C, they would be treated as multiple unique patients (one per site). This is a known limitation of the dataset, but it is unknown if this scenario occurred in any of the cohorts in the present analysis. Additionally, the selected cohorts are not mutually exclusive. A patient with diabetes type 1 and controlled hypertension will be counted in both cohorts. This is again a known limitation of our dataset, but of low consequence since this study does not aim to look at interaction of diseases but only investigates proportions across any of the selected diseases.

### Site inclusion criteria

The N3C dataset contains positive cases and controls in the ratio of 1:2 matched by age, sex, and race (see Patient Cohort Selection). Due to this inclusion criteria, the analyses in this study cannot accurately estimate the absolute incidence or prevalence of COVID-19 in selected disease cohorts. Additionally, there are two factors that introduce observation bias into the dataset: 1) the site contributing data, and 2) age of the patient (which is considered a contributing factor to COVID-19 susceptibility and disease severity and is also an important factor in the development of signs and symptoms related to NF1, NF2, or SWN, and thus having a recorded diagnosis of one of these conditions).

Contributing sites may also introduce confounding factors into disease severity metrics, like the variability of criteria applied for use of various interventions, testing or procedures. For example, there may be different indications for intubation at various contributing health care sites or there may be variable availability of a given resource or procedure across sites. Additionally, since the clinical data provided by contributing sites varies in granularity, some sites have systematic missingness of some variables. To control for these differences and missingness between sites and data partners contributing to our selected cohorts, we only included patient data from sites that contributed NF1, NF2, or SWN patients. To achieve this exclusion, first, the unique “data partner ids” (correlating to healthcare sites) in the NF1, NF2, and SWN cohorts were noted. These data partner ids were then used to filter all the other cohorts so that only patients contributed by sites that contributed NF1, NF2, or SWN patients were selected for inclusion.

### Age adjustment for selected disease cohorts

Age-related differences between the cohorts were adjusted by stratifying the cohorts into 10-year age bins. Each stratum was then weighted using the age-adjusted rate (“aarate”) formula based on US standard population (US census 2000)^10,11^. Each age-adjusted disease cohort comprised of the ages x through y and was calculated using the following formula:

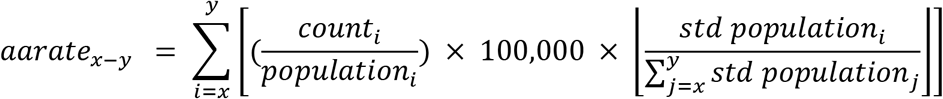

Age-adjusted counts of positive cases, severe outcomes, or invasive ventilation for each cohort are available as supplemental tables. More details of the steps involved in the calculations of age-adjusted counts are available in the supplementary tables 11,12, and 13.

### Bootstrap analyses

Bootstrapping is a standard resampling technique that allows direct visualization of confidence intervals of a statistic when 1) no assumption about the parametric or non-parametric distribution of the statistic can be made and/or 2) an experiment cannot be repeated multiple times to generate the statistic again.^12^ In this study we compared the age-adjusted proportions of positive cases in different selected cohorts using a standard z-test. We then used bootstrap analysis to estimate confidence intervals of the p-value statistics derived the standard z-test. Three main groups of comparisons were assessed: *NF1 vs all other study cohorts, DM1 vs all other study cohorts*, and *NF1 vs all other “rare” cohorts pooled (TS, FX, MCC, AML, NF2, SWN)*. For the *NF1 vs all* comparison, a test vector was populated with the age-stratified proportions from the NF1 cohort. A comparison vector was populated with the age-stratified proportions of all the other cohorts. A Shapiro-Wilk test (base R v3.6.3 `shapiro.test` function) was used to test the normality of the distributions of the age-stratified proportions for each cohort (NF1 cohort: p-value = 0.165, Shapiro-Wilk test). The age stratified proportions were compared to estimate the p-value of the real observations (“real p-value”) (using the `z.test` function from the package BSDA v1.2.0). Then, the age-adjusted proportions in the test and comparison vectors were resampled 10,000 times to produce 10,000 possible combinations of age-adjusted proportions (using the `resample` function from the `gdata v2.18.0` R package). For each of these “resampled cohorts”, a z-test was performed to estimate the distribution of possible p-values generated from the observed proportions. This distribution of p-values generated through bootstrap presents the confidence intervals for the observed “real” p-value. If the “real” p-value was not significantly different (Wilcoxon rank sum test) from the distribution of various p-values generated in the bootstrap and was less than p=0.05, the real p-value was unlikely to occur by chance, and the cohorts in the comparison were considered significantly different. A similar approach was taken for all other comparisons of proportions, except that a non-parametric Wilcoxon rank sum test (base R v3.6.3 `wilcox.test function)` was used for comparisons for severe outcomes and invasive ventilation (due to non-normal distribution in all cohorts). All real p-values were adjusted to correct for the number of overlapping comparisons for each disease using Benjamini-Hochberg method. The distributions of bootstrapped p-values were visualized using R `ggplot2 v3.3.2`. Similar analyses were also done for NF2 and SWN cohorts.

### Confidence interval calculations

All comparisons were tested using 95% confidence interval as default. In some bootstrap analysis comparisons, the skew in the distribution of values did not allow confidence interval calculations at 95% (as the difference between α achieved from the distribution and α_target_ was greater than α_target_/2, where α_target_ = 0.05). In such cases, the highest confidence interval that was able to be calculated is reported (60%). It should be noted that a 60% confidence interval is more likely to reject the null hypothesis as compared to a 95% CI. In this study confidence intervals were calculated and are reported at 95%, any comparisons with 60% CI have been explicitly noted in the tables.

## RESULTS

### Demographics of the NF1, NF2, and SWN cohorts are comparable to those of other cohorts in N3C

From 6.4 million patients present in N3C Data enclave (v3.3, July 2021), we selected cohorts of rare (NF1: 2,501, NF2: 665, SWN: 762, TS: 861, AML: 9,844, FX: 98, MCC: 648) and non-rare diseases (non-NF1: 5.6 million, non-NF2: 5.6 million, non-SWN: 5.6 million, DM1: 66,234, HYP: 1.6 million) using concept sets of EHR diagnosis codes (see Methods, Table 1). The cohort of fragile X syndrome (FX) was the smallest among all selected cohorts. Other rare disease cohorts were comparable in number to each other while considerably smaller than the non-rare disease cohorts, as expected. The occurrence of NF1, NF2, and SWN patients in the N3C data (NF1: 0.0004 of total N3C patients, NF2: 0.0001 of total N3C patients, SWN: 0.0001 of total N3C patients) was found to be higher than the expected population prevalence of these diseases (NF1: 0.0002, NF2: 0.00002, SWN: 0.000008 approximately), indicating that the N3C dataset may not represent a random sample of the general population (see Supplemental Methods).

**Table 1:** Table showing the number of unique persons in each of the selected cohorts. (NF1 : Neurofibromatosis type 1, TS: tuberous sclerosis, AML: acute myeloid leukemia, FX: fragile-X syndrome, MCC: Merkel cell carcinoma, Non-NF1: general population without NF1, Non-NF2: general population without NF2, Non-SWN: general population without SWN, DM1: diabetes mellitus type-1, HYP: controlled hypertension)

|  | NF1 | NF2 | SWN | TS | AML | FX | MCC | Non-NF1 | Non-NF2 | Non-SWN | DM1 | HYP |
| --- | --- | --- | --- | --- | --- | --- | --- | --- | --- | --- | --- | --- |
| Site-adjusted Cohort Size | 2,501 | 665 | 762 | 861 | 9,844 | 98 | 648 | 5,577,737 | 5,579,215 | 5,579,146 | 66,234 | 1,664,134 |
| Median Age (years) | 28 | 49 | 60 | 23 | 62 | 28 | 75 | 46 | 46 | 46 | 50 | 63 |

The median ages of the NF1, FX, and TS cohorts were substantially lower than the non-NF1 population or the non-rare disease populations (Table 1), while the median ages of the NF2 and SWN cohorts were more comparable to the non-NF population. This suggests that age-related adjustments are necessary before comparing the cohorts. The selected NF1, NF2, and SWN cohorts were similar to the non-NF1, non-NF2, or non-SWN N3C population with regards to race, with a majority of white but a substantial representation from the black or African-American race (Figure 1A, Supplemental Table 10). The NF1, NF2, and SWN cohorts and the general population cohorts had similar distributions of male and female patients (Figure 1B, Supplemental Table 10).

**Figure 1.**
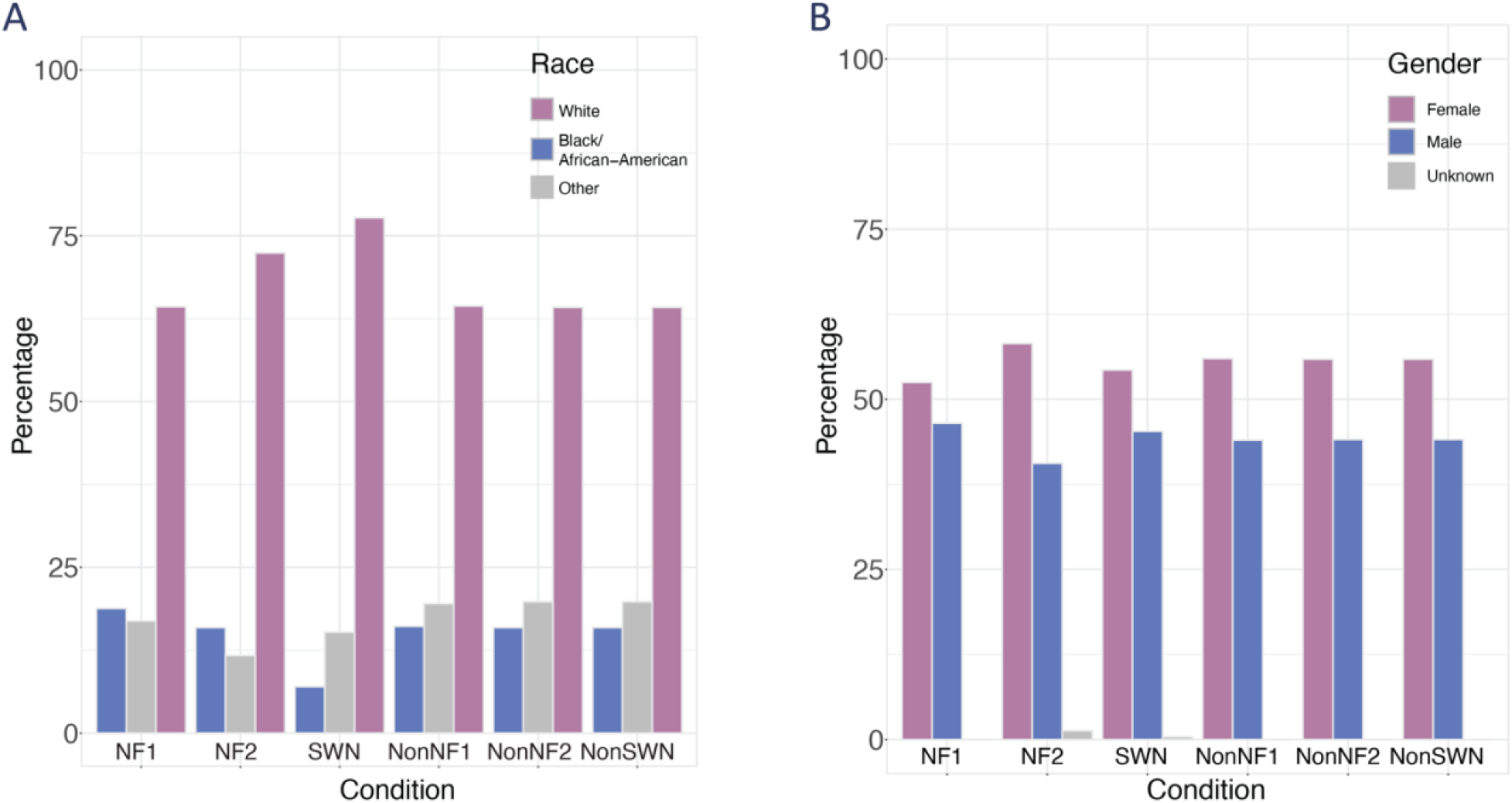
Demographics of selected cohorts in N3C. (A) Bar-plot showing percentage of unique persons that identify as White, Black, or Other races in the selected cohorts. (NF1 : Neurofibromatosis type 1, TS: tuberous sclerosis, AML: acute myeloid leukemia, FX: fragile-X syndrome, MCC: Merkel cell carcinoma, Non-NF1: general population without NF1, Non-NF2: general population without NF2, Non-SWN: general population without SWN) (C) Bar-plot showing percentage of unique persons identifying as Male or Female in NF1, NF2, SWN, Non-NF1, Non-NF2, and Non-SWN cohorts.

### Age-adjusted proportion of SARS-CoV-2 positive cases in NF1, NF2, or SWN is not greater than other selected diseases

To test whether SARS-CoV-2 affected the NF1 population differently than other populations, we compared the age-adjusted proportions of positive cases (SARS-CoV-2 positive and/or COVID-19 patients) in the NF1 cohort with that of the non-NF1 population, other rare diseases, and selected non-rare disease cohorts (Figure 2, Supplemental Table 11, Table 2-3). We made similar comparisons for NF2 and SWN. The proportions of positive cases in the NF1, NF2, and SWN cohorts were low compared to those of other groups (NF1: 14.5%, NF2: 13.8%, SWN: 13.7%, DM1: 24.0%, Table 2). Comparison of the proportions of positive cases in the NF1 cohort with all the other cohorts showed that the real p-value (red dashed line) was less than 0.05 (Figure 2A, Table 3). Moreover, the estimated confidence in the observed p-value through bootstrap analysis suggests that this was not observed by chance but fell within the distribution of various possible p-values that can be generated from the observed proportions (z-test p-value = 0.0028, Benjamini-Hochberg [BH] adjusted p-value = 0.008 and falls within bootstrap distribution; Wilcoxon rank sum test p-value = 0.5, Table 3). This suggests that the proportion of positive cases in the NF1 N3C cohort was not higher than the non-NF1 N3C cohort. Similarly, Figures 2D and Table 3 show that the NF2 N3C cohort also had a significantly lower proportion of positive cases than the non-NF2 N3C cohort (z-test p-value = 3.9×10^−5^, Benjamini-Hochberg [BH] adjusted p-value = 1.2×10^−4^, and falls within bootstrap distribution: Wilcoxon rank sum test p-value = 0.5). The age-adjusted proportions of positive cases in the SWN cohort, however, did not differ significantly from the other cohorts (z-test p-value = 0.05, Benjamini-Hochberg [BH] adjusted p-value = 0.16, and falls within bootstrap distribution: Wilcoxon rank sum test p-value = 1.0, Figure 2G, Table 3).

**Figure 2.**
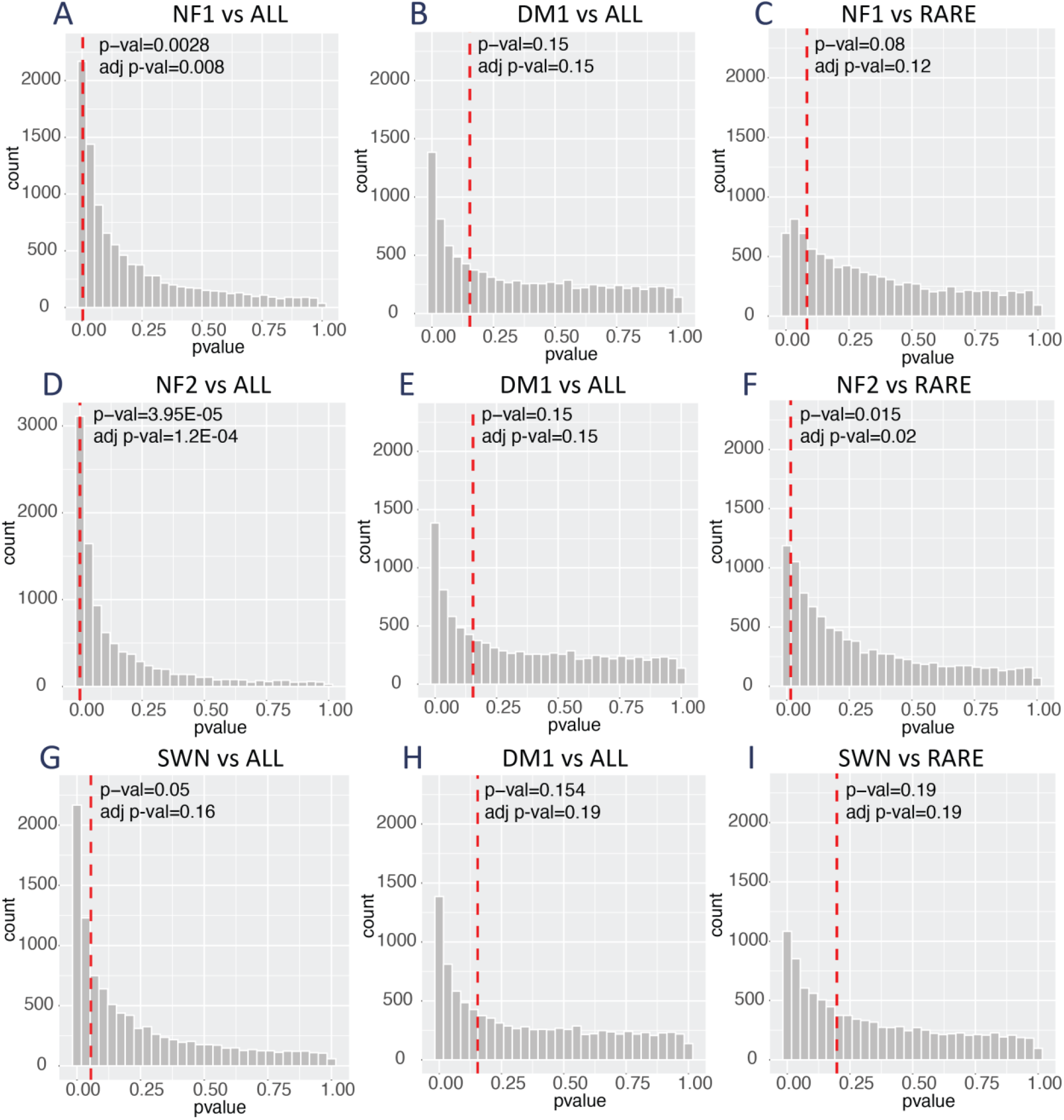
Comparison of age-adjusted proportions of positive cases in selected cohorts. Selected cohorts include NF1: Neurofibromatosis type 1, TS: tuberous sclerosis, AML: acute myeloid leukemia, FX: fragile-X syndrome, MCC: Merkel cell carcinoma, Non-NF1/Non-NF2/Non-SWN: general population, DM1: diabetes mellitus type 1, HYP: controlled hypertension (A-C) Results of the bootstrap analysis for p-value of comparisons between NF1 and all, DM1 vs. all, NF1 vs. rare disease cohorts. (D-F) Results of the bootstrap analysis for comparisons between NF2 and all, DM1 vs all, NF2 vs rare disease cohorts. (G-I) Results of the bootstrap analysis for comparisons of p-values between SWN and all, DM1 vs. all, SWN vs rare disease cohorts. The red dashed line represents the p-value obtained from the real observations. The specific and adjusted value for each comparison is noted in the plot inset. The grey bars show a histogram of all possible p-values obtained through 10,000 iterations of bootstrap analysis.

**Table 2:** Table showing the age-adjusted counts and percentages of positive cases in each cohort. (NF1: Neurofibromatosis type 1, TS: tuberous sclerosis, AML: acute myeloid leukemia, FX: fragile-X syndrome, MCC: Merkel cell carcinoma, Non-NF1: general population, DM1: diabetes mellitus type 1, HYP: controlled hypertension)

|  | NF1 | NF2 | SWN | TS | AML | FX | MCC | Non-NF1 | Non-NF2 | Non-SWN | DM1 | HYP |
| --- | --- | --- | --- | --- | --- | --- | --- | --- | --- | --- | --- | --- |
| Age-adjusted counts of positive cases per 100,000 US standard population | 14,496 | 13,782 | 13,738 | 19,346 | 18,135 | 28,105 | 19823 | 28,503 | 28,498 | 28,498 | 24,022 | 24,949 |
| Approximate percentage of positive cases | 14.5 | 13.8 | 13.7 | 19.3 | 18.1 | 28.1 | 19.8 | 28.5 | 28.5 | 28.5 | 24.0 | 24.9 |

**Table 3:**
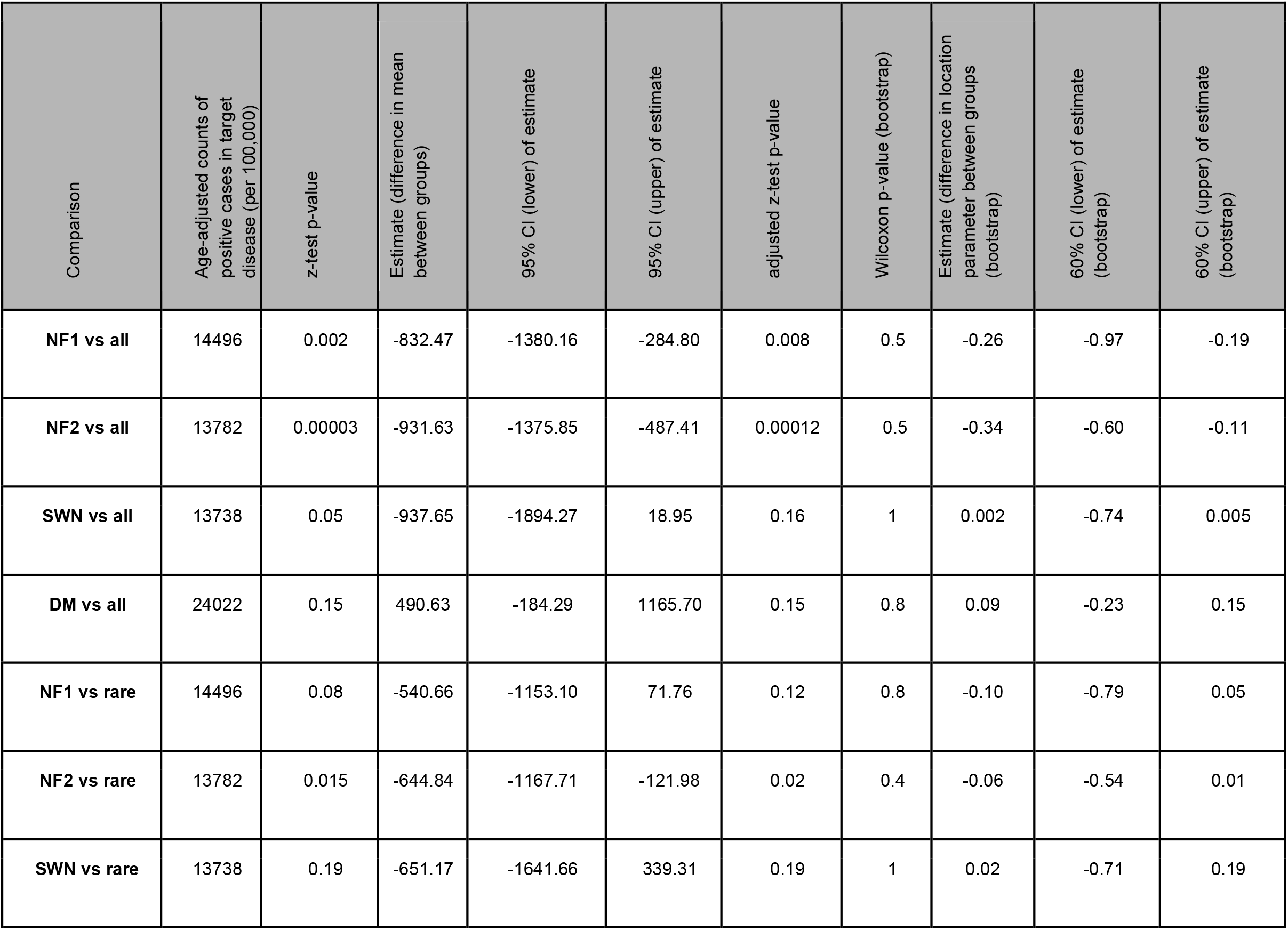
Table showing age-adjusted counts and p-values of all comparisons of positive cases in selected cohorts (as shown in Figure 2 A-F). Confidence interval is abbreviated as CI in the table. In the bootstrap analysis, the z-test p-value was compared to a distribution of bootstrapped p-values using the non-parametric Wilcoxon rank sum test. In this test, the null hypothesis is that the two distributions differ by a location shift of µand the alternative hypothesis is that they differ by a location shift other than µ. The “estimate” of this non-parametric test is equal to the difference in µwhich in the present case has negative values due to the direction of the location shift. The confidence intervals reflect the range of µand has negative values. The skew in the distribution of bootstrapped p-values did not allow confidence interval calculations at 95% (as the difference between α achieved from the distribution and α_target_ was greater than α_target_/2, where α_target_ = 0.05), but enabled 60% CI estimation. A 60% confidence interval is more likely to reject the null hypothesis as compared to a 95% CI.

| Comparison | Age-adjusted counts of positive cases in target disease (per 100,000) | z-test p-value | Estimate (difference in mean between groups) | 95% CI (lower) of estimate | 95% CI (upper) of estimate | adjusted z-test p-value | Wilcoxon p-value (bootstrap) | Estimate (difference in location parameter between groups (bootstrap) | 60% CI (lower) of estimate (bootstrap) | 60% CI (upper) of estimate (bootstrap) |
| --- | --- | --- | --- | --- | --- | --- | --- | --- | --- | --- |
| **NF1 vs all** | 14496 | 0.002 | -832.47 | -1380.16 | -284.80 | 0.008 | 0.5 | -0.26 | -0.97 | -0.19 |
| **NF2 vs all** | 13782 | 0.00003 | -931.63 | -1375.85 | -487.41 | 0.00012 | 0.5 | -0.34 | -0.60 | -0.11 |
| **SWN vs all** | 13738 | 0.05 | -937.65 | -1894.27 | 18.95 | 0.16 | 1 | 0.002 | -0.74 | 0.005 |
| **DM vs all** | 24022 | 0.15 | 490.63 | -184.29 | 1165.70 | 0.15 | 0.8 | 0.09 | -0.23 | 0.15 |
| **NF1 vs rare** | 14496 | 0.08 | -540.66 | -1153.10 | 71.76 | 0.12 | 0.8 | -0.10 | -0.79 | 0.05 |
| **NF2 vs rare** | 13782 | 0.015 | -644.84 | -1167.71 | -121.98 | 0.02 | 0.4 | -0.06 | -0.54 | 0.01 |
| **SWN vs rare** | 13738 | 0.19 | -651.17 | -1641.66 | 339.31 | 0.19 | 1 | 0.02 | -0.71 | 0.19 |

In contrast to NF1 and NF2, the proportion of positive cases observed in a non-rare disease like diabetes mellitus type 1 (DM1) were not significantly different from that of the rest of the cohorts (z-test p-value = 0.15, BH adjusted p-value = 0.15, and falls within bootstrap distribution: Wilcoxon rank sum test p-value = 0.8, Figures 2B, 2E, 2H, Table 3).

We further compared the age-adjusted proportions of positive cases noted in the NF1 cohort with the other selected rare diseases to test whether all rare diseases considered in this study tended to have lower proportions of positive cases or if this was unique to NF1. The proportion of SARS-CoV-2 positive or COVID-19 patients in the NF1 cohort was not significantly different from all the other rare disease cohorts (NF2, SWN, TS, MCC, AML, FX) considered as a single group (z-test p-value = 0.08, BH adjusted p-value = 0.12, falls within bootstrap distribution: Wilcoxon rank sum test p-value = 1, Figure 2C, Table 3). A similar trend was noted for the SWN cohort (z-test p-value = 0.19, BH adjusted p-value = 0.19, falls within bootstrap distribution: Wilcoxon rank sum test p-value = 1, Figure 2I, Table 3). Interestingly, the age-adjusted proportions of positive cases in the NF2 cohort were significantly lower compared to other rare disease cohorts (z-test p-value = 0.015, BH adjusted p-value = 0.02, falls within bootstrap distribution: Wilcoxon rank sum test p-value = 0.4, Figure 2F, Table 3).

Interpreting the above results conservatively, the proportions of positive cases in the N3C NF1, NF2, and SWN populations were no greater than expected for rare or non-rare diseases examined in this study.

### Age-adjusted proportion of severe outcomes in NF1, NF2, and SWN was not greater than that of other diseases

Though the positive cases did not appear to be more frequent in people with NF1, NF2, and SWN versus those without these diseases, it is possible that the severity of COVID-19 in positive cases with NF1, NF2, or SWN is different from that of the other diseases. In the N3C cohort, there were no patients with NF2, SWN, FX or TS that had reported severe outcomes; thus, we were unable to statistically evaluate the prevalence of severe outcomes in NF2 or SWN cohorts. We evaluated the severity of COVID-19 manifestations in the NF1 cohort and compared that to other selected cohorts. N3C has made extensive efforts to capture the severity of disease incorporating information from EHRs such as hospitalization, invasive ventilation, extracorporeal membrane oxygenation, hospice, and death.^8^ We examined patient severity scores built on these parameters in our selected cohorts to estimate the severity of COVID-19.

We first identified the patients in the previously examined disease cohorts with highest documented severity^13^ (N3C severity type Severe, i.e., WHO severity 7-9, and Mortality/Hospice, i.e., WHO severity 10). We then compared the age-adjusted proportions of patients with these severity types (henceforth referred to as “severe outcomes”) among positive cases in each cohort (Figure 3A-C, Supplemental Table 12). We found that the proportion of severe outcomes in the NF1 cohort was not significantly different when compared to all other cohorts (Wilcoxon rank sum test, observed p-value = 0.56, BH adjusted p-value = 0.56, falls within bootstrap distribution: p-value = 0.8) (Figure 3A, Table 4). In contrast, we found that the DM1 cohort had higher proportions of patients with severe outcomes compared to the other selected cohorts (Wilcoxon rank sum test, observed p-value = 0.003, BH adjusted p-value = 0.009, falls within bootstrap distribution: p-value = 1.0) (Figure 3B, Table 4). This finding is consistent with the now established association between diabetes and severity of COVID-19 outcomes^14,15^. The proportion of patients with severe outcomes in the NF1 cohort was not significantly higher than the other rare disease cohorts examined, suggesting no clear relationship between NF1 and severe outcome from COVID-19 infection/disease (Wilcoxon rank sum test, observed p-value = 0.04, BH adjusted p-value = 0.06, falls within bootstrap distribution: p-value = 0.28) (Figure 3C, Table 4).

**Figure 3.**
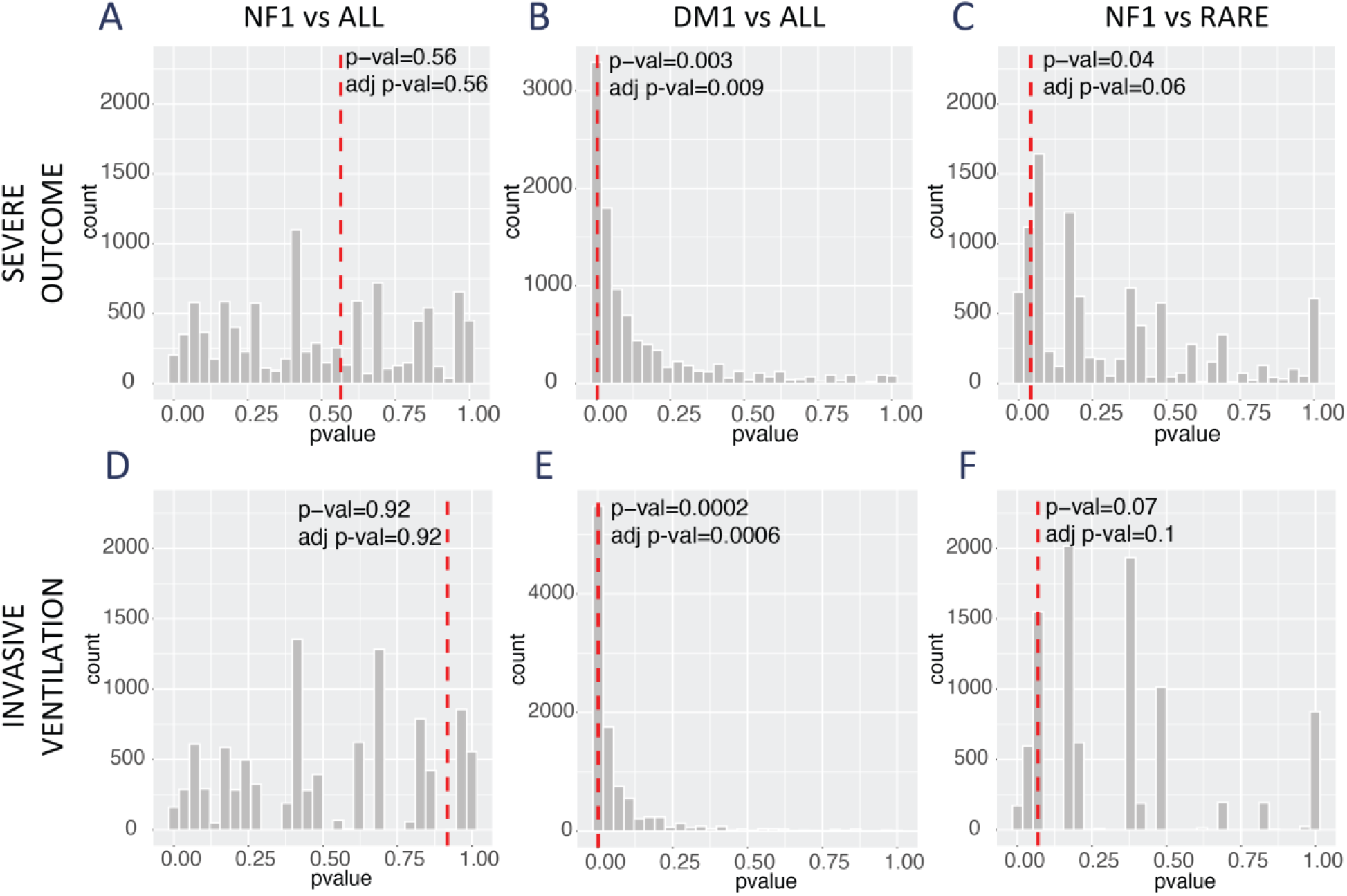
Comparison of age-adjusted proportions of severe outcomes and invasive ventilation in selected cohorts. (A-C) Results of the bootstrap analysis for comparisons of proportions that experienced severe outcomes between NF1 vs all, DM1 vs all, and NF1 vs rare disease cohorts. The red dashed line represents the p-value obtained from the real observations. The specific value for each comparison is noted in the plot inset. The grey bars show a histogram of all possible p-values obtained through 10,000 iterations of bootstrap analysis. (D-F) Results of the bootstrap analysis for comparisons of proportions that required invasive ventilation between NF1 vs all, DM1 vs all, and NF1 vs rare disease cohorts. The red dashed line represents the p-value obtained from the real observations. The specific value for each comparison is noted in the plot inset. The grey bars show a histogram of all possible p-values obtained through 10,000 iterations of bootstrap analysis.

**Table 4:** Table showing age-adjusted counts, confidence intervals (CI), and p-values of all comparisons of severe outcomes in various selected cohorts. In some cases, the skew in the distribution of bootstrapped p-values did not allow confidence interval calculations at 95% CI (as the difference between α achieved from the distribution and α_target_ was greater than α_target_/2, where α_target_ = 0.05). Confidence interval calculations at the 60% level are reported for these comparisons (indicated by *). The confidence limits without an asterisk denote 95% CI. A 60% confidence interval is more likely to reject the null hypothesis as compared to a 95% CI.

| Comparison | Age-adjusted counts of severe outcomes in target disease (per 100,000) | Wilcoxon p-value | Estimate (difference in location parameter between groups) | 95% CI (lower) of estimate | 95% CI (upper) of estimate | adjusted Wilcoxon p-value | Wilcoxon p-value (bootstrap) | Estimate (difference in location parameter between groups) (bootstrap) | CI (lower) of estimate (bootstrap) | CI (upper) of estimate (bootstrap) |
| --- | --- | --- | --- | --- | --- | --- | --- | --- | --- | --- |
| **NF1 vs all** | 887 | 0.56 | 0.0000016 | -0.00006 | 204.70 | 0.56 | 0.8 | -0.18 | -0.43* | -0.04* |
| **DM vs all** | 534 | 0.003 | 42.10 | 28.05 | 64.43 | 0.009 | 1 | -0.07 | -0.36* | 0.002* |
| **NF1 vs rare** | 887 | 0.04 | 0.000002 | -0.000019 | 204.70 | 0.06 | 0.28 | -0.04 | -0.34 | -0.034 |

We also examined the proportions of positive cases in the NF1 cohort who received invasive ventilation (Figure 3D-F, Table 5, Supplemental Table 13). The proportions requiring invasive ventilation among the positive cases in the NF1 cohort were not significantly different from the other cohorts (Wilcoxon rank sum test, observed p-value = 0.91, BH adjusted p-value = 0.92, falls within bootstrap distribution: p-value = 1.0, Figure 3D, F, & Table 5). In contrast, the DM1 cohort appears to have more invasive ventilation (Wilcoxon rank sum test, observed p-value = 0.0002, BH adjusted p-value = 0.0006) (Figure 3E, Table 5). The median length of hospital stays for the patients with NF1 who had severe outcomes was not substantially different than other cohorts (NF1: 10 days, AML: 9 days, MCC: 23 days, Non-NF1: 11 days, DM1: 13 days, HYP: 11 days, TS: not determined., FX: not determined., NF2: not determined., SWN: not determined.; Wilcoxon rank sum test p-value = 0.55, Supplemental Table 14). Thus, our findings suggest that the proportion of positive cases in the NF1 cohort experiencing severe outcomes was not significantly greater than that in either non-rare or rare disease cohorts.

**Table 5:**
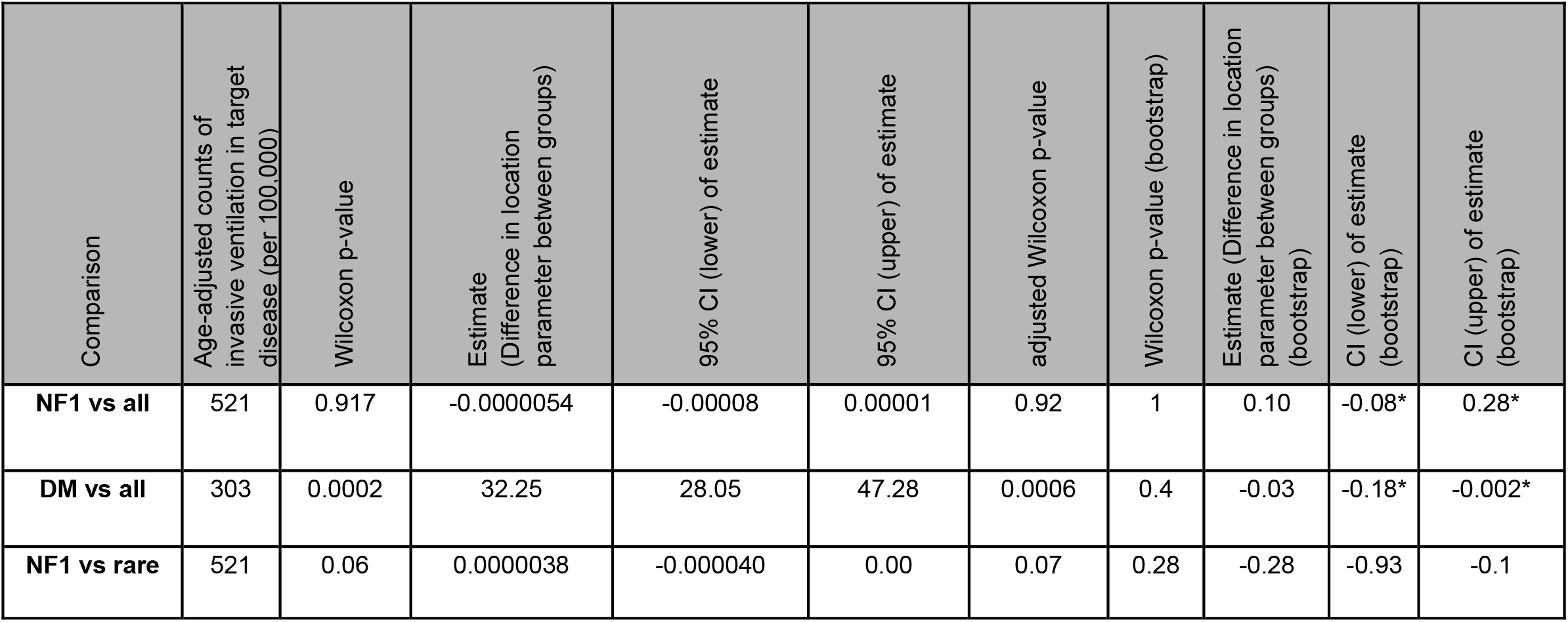
Table of age-adjusted counts, confidence intervals (CI) and p-values of all comparisons of invasive ventilation recorded in selected cohorts. In some cases, the skew in the distribution of bootstrapped p-values did not allow confidence interval calculations at 95% CI (as the difference between α achieved from the distribution and α_target_ was greater than α_target_/2, where α_target_ = 0.05). Confidence interval calculations at the 60% level are reported for these comparisons (indicated by *). The confidence limits without an asterisk denote 95% CI. A 60% confidence interval is more likely to reject the null hypothesis as compared to a 95% CI.

| Comparison | Age-adjusted counts of invasive ventilation in target disease (per 100,000) | Wilcoxon p-value | Estimate (Difference in location parameter between groups) | 95% CI (lower) of estimate | 95% CI (upper) of estimate | adjusted Wilcoxon p-value | Wilcoxon p-value (bootstrap) | Estimate (Difference in location parameter between groups) (bootstrap) | CI (lower) of estimate (bootstrap) | CI (upper) of estimate (bootstrap) |
| --- | --- | --- | --- | --- | --- | --- | --- | --- | --- | --- |
| **NF1 vs all** | 521 | 0.917 | -0.0000054 | -0.00008 | 0.00001 | 0.92 | 1 | 0.10 | -0.08* | 0.28* |
| **DM vs all** | 303 | 0.0002 | 32.25 | 28.05 | 47.28 | 0.0006 | 0.4 | -0.03 | -0.18* | -0.002* |
| **NF1 vs rare** | 521 | 0.06 | 0.0000038 | -0.000040 | 0.00 | 0.07 | 0.28 | -0.28 | -0.93 | -0.1 |

## DISCUSSION

In this study we examined the EHR data present in the N3C Data Enclave to determine the burden of SARS-CoV-2 in people with NF1, NF2, and SWN. Our data give no evidence for patients with NF being at higher risk of SARS-CoV-2 infection or COVID-19 when adjusted for age and site. Our findings suggest that the proportion of positive cases and severe outcomes among patients with NF1, NF2, and SWN in the N3C Data Enclave was not higher than other selected rare and non-rare diseases.

The N3C is the largest centralized and harmonized EHR repository of a representative COVID-19 cohort in the United States to date^8^, well suited for research on COVID-19 related outcomes. While it is an extensive collection of EHR data, various limitations exist in using this dataset to determine SARS-CoV-2 and COVID-19-related risks for the global population. For example, we observed a greater prevalence of NF1 patients in the N3C as compared to population prevalence estimates, indicating that this dataset may not represent the general population due to its specific data acquisition protocols. As with any multi-site combination of EHR data, there may also be site-related differences in clinical measures due to variations in clinical practice and medical record documentation. Any biases affecting analyses of care patterns or outcomes due to geographical, regional, cultural, or other differences between institutions remained unassessed due to anonymized coding of the N3C data contributing institutions in the de-identified dataset. Furthermore, clinical coding of patients with NF1, NF2, SWN or other rare diseases may be incomplete in EHRs, i.e. the codes used to define the study cohorts (Supplemental Tables 1-9) within N3C could have missed disease-positive patients without appropriately recorded diagnostic codes. Lastly, the SARS-CoV-2 testing rate, diagnosis and treatment of COVID-19, access to clinical care in various sites and various disease populations may vary significantly.

Additionally, there are various considerations specific to the data deposited in the N3C data enclave. Due to N3C’s phenotype acquisition design, the patients included in the Data Enclave were matched by demographics but not disease type. This matching strategy suggests that the N3C may not provide an accurate proportion of the SARS-CoV-2 negative/non-COVID-19 population for different disease cohorts, limiting the comparisons to those across different disease cohorts within the N3C. It is also important to note that the definition of “SARS-CoV-2-positive or COVID-19 patients” in this study is subject to limitations such as potential false positives, false negatives, and untested asymptomatic individuals. This database also lacks records of individuals who did not have a clinical encounter at N3C contributing sites due to being suspected positive or asymptomatic, which could also bias the results. Furthermore, the present study only focuses on the acute data related to SARS-CoV-2 infections and associated critical care usage. Future analyses evaluating additional patient covariates known to impact SARS-CoV-2 outcomes, such as pregnancy ^16^, or long-COVID data may help us refine the results of this study. Finally, sample sizes for certain diseases (e.g. FX), makes interpretation of the results for these diseases challenging. Additional data for the cohorts may also allow for higher confidence interval estimates for comparisons in the study where the present data only enabled 60% confidence interval calculations.

Despite these limitations, we recapitulated well-established associations between COVID-19 and DM1 (one of our control cohorts), suggesting that our methods can identify underlying patterns of SARS-CoV-2 risk and severity in a common disease.^14,17,18^ Similar to previous observations, our findings suggest that while the proportion of DM1 patients found to be positive cases is not greater than the general population (Figure 2C), the proportion of positive cases in the DM1 cohort experiencing severe outcomes was higher compared to the rest of the comparison cohorts (Figure 3B, E, Table 2). These observations in our analysis are reassuring and indicate that our analytical methods may be tolerant of the inherent biases and limitations of the N3C dataset and EHR data while identifying robust patterns for selected cohorts. In the future, additional studies (e.g., case-control health surveys, mobile health studies) may allow more accurate determination of the prevalence of COVID-19 and its impact on health in people with NF1, NF2, and SWN. Future studies should also evaluate whether people with rare diseases exhibit cautious behavior or stronger adherence to social distancing protocols contributing to lower SARS-CoV-2 infection rates.

This study leverages a new and unique dataset and overcomes various statistical challenges to assess COVID-19 burden and severity in a rare disease population. We anticipate that the strategies used in this study can be easily extended to examine other rare diseases of interest using the N3C dataset, thus serving as a roadmap for future work. Using these methods, we discovered that people with NF1, NF2, and SWN do not appear to be at a greater risk of becoming positive cases or developing severe complications of COVID-19 compared to other rare or non-rare diseases. These findings suggest that while no elevated risk was noted as per the composition of N3C patient population in July 2021, it is important for people with NF to follow COVID-19-related public health measures, vaccination guidelines, and recommendations from NF specialists.^19^

## Data Availability

All data analyzed in this study are available online at the N3C Data Enclave (https://covid.cd2h.org/enclave). Access to data requires approval from the N3C data access committee. No new data was produced in this study. All the R code used for the analyses in this study is available on Github (https://github.com/jaybee84/NF-COVID-response). 

## CODE AND DATA AVAILABILITY

Data are available in the N3C Data Enclave (https://covid.cd2h.org/enclave). All the R code used in the analyses in this study is available on Github.

## ACKNOWLEDGEMENTS

Authorship was determined using ICMJE recommendations. We thank Dr. Tim Bergquist, Yao Yan, and Dr. Justin Guinney at Sage Bionetworks for helpful input during the design of this project. We additionally thank Dr. Harold Lehmann (Johns Hopkins School of Medicine), and Dr. Kenneth Wilkins (NIDDK) for helpful and insightful discussions regarding the statistical analysis strategies described in this study.

## N3C Attribution

The analyses described in this publication were conducted with data or tools accessed through the NCATS N3C Data Enclave https://covid.cd2h.org and N3C Attribution & Publication Policy v 1.2-2020-08-25b supported by NCATS U24 TR002306. This research was possible because of the patients whose information is included within the data and the organizations (https://ncats.nih.gov/n3c/resources/data-contribution/data-transfer-agreement-signatories) and scientists who have contributed to the on-going development of this community resource [https://doi.org/10.1093/jamia/ocaa196].”

### IRB

The N3C data transfer to NCATS is performed under a Johns Hopkins University Reliance Protocol # IRB00249128 or individual site agreements with NIH. The N3C Data Enclave is managed under the authority of the NIH; information can be found at https://ncats.nih.gov/n3c/resources.

### Individual Acknowledgements for Core Contributors

We gratefully acknowledge contributions from the following N3C core teams

(Asterisks indicate leads) • Principal Investigators: Melissa A. Haendel*, Christopher G. Chute*, Kenneth R. Gersing, Anita Walden

- Workstream, subgroup and administrative leaders: Melissa A. Haendel*, Tellen D. Bennett, Christopher G. Chute, David A. Eichmann, Justin Guinney, Warren A. Kibbe, Hongfang Liu, Philip R.O. Payne, Emily R. Pfaff, Peter N. Robinson, Joel H. Saltz, Heidi Spratt, Justin Starren, Christine Suver, Adam B. Wilcox, Andrew E. Williams, Chunlei Wu
- Key liaisons at data partner sites
- Regulatory staff at data partner sites
- Individuals at the sites who are responsible for creating the datasets and submitting data to N3C • Data Ingest and Harmonization Team: Christopher G. Chute*, Emily R. Pfaff*, Davera Gabriel, Stephanie S. Hong, Kristin Kostka, Harold P. Lehmann, Richard A. Moffitt, Michele Morris, Matvey B. Palchuk, Xiaohan Tanner Zhang, Richard L. Zhu
- Phenotype Team (Individuals who create the scripts that the sites use to submit their data, based on the COVID and Long COVID definitions): Emily R. Pfaff*, Benjamin Amor, Mark M. Bissell, Marshall Clark, Andrew T. Girvin, Stephanie S. Hong, Kristin Kostka, Adam M. Lee, Robert T. Miller, Michele Morris, Matvey B. Palchuk, Kellie M. Walters
- Project Management and Operations Team: Anita Walden*, Yooree Chae, Connor Cook, Alexandra Dest, Racquel R. Dietz, Thomas Dillon, Patricia A. Francis, Rafael Fuentes, Alexis Graves, Julie A. McMurry, Andrew J. Neumann, Shawn T. O’Neil, Usman Sheikh, Andréa M. Volz, Elizabeth Zampino
- Partners from NIH and other federal agencies: Christopher P. Austin*, Kenneth R. Gersing*, Samuel Bozzette, Mariam Deacy, Nicole Garbarini, Michael G. Kurilla, Sam G. Michael, Joni L. Rutter, Meredith Temple-O’Connor
- Analytics Team (Individuals who build the Enclave infrastructure, help create codesets, variables, and help Domain Teams and project teams with their datasets): Benjamin Amor*, Mark M. Bissell, Katie Rebecca Bradwell, Andrew T. Girvin, Amin Manna, Nabeel Qureshi
- Publication Committee Management Team: Mary Morrison Saltz*, Christine Suver*, Christopher G. Chute, Melissa A. Haendel, Julie A. McMurry, Andréa M. Volz, Anita Walden
- Publication Committee Review Team: Carolyn Bramante, Jeremy Richard Harper, Wenndy Hernandez, Farrukh M Koraishy, Federico Mariona, Amit Saha, Satyanarayana Vedula

### Data Partners with Released Data

Stony Brook University — U24TR002306 • University of Oklahoma Health Sciences Center — U54GM104938: Oklahoma Clinical and Translational Science Institute (OCTSI) • West Virginia University — U54GM104942: West Virginia Clinical and Translational Science Institute (WVCTSI) • University of Mississippi Medical Center — U54GM115428: Mississippi Center for Clinical and Translational Research (CCTR) • University of Nebraska Medical Center — U54GM115458: Great Plains IDeA-Clinical & Translational Research • Maine Medical Center U54GM115516: Northern New England Clinical & Translational Research (NNE-CTR) Network • Wake Forest University Health Sciences — UL1TR001420: Wake Forest Clinical and Translational Science Institute • Northwestern University at Chicago — UL1TR001422: Northwestern University Clinical and Translational Science Institute (NUCATS) • University of Cincinnati — UL1TR001425: Center for Clinical and Translational Science and Training • The University of Texas Medical Branch at Galveston — UL1TR001439: The Institute for Translational Sciences • Medical University of South Carolina — UL1TR001450: South Carolina Clinical & Translational Research Institute (SCTR) • University of Massachusetts Medical School Worcester — UL1TR001453: The UMass Center for Clinical and Translational Science (UMCCTS) • University of Southern California — UL1TR001855: The Southern California Clinical and Translational Science Institute (SC CTSI) • Columbia University Irving Medical Center — UL1TR001873: Irving Institute for Clinical and Translational Research • George Washington Children’s Research Institute — UL1TR001876: Clinical and Translational Science Institute at Children’s National (CTSA-CN) • University of Kentucky — UL1TR001998: UK Center for Clinical and Translational Science • University of Rochester — UL1TR002001: UR Clinical & Translational Science Institute • University of Illinois at Chicago — UL1TR002003: UIC Center for Clinical and Translational Science • Penn State Health Milton S. Hershey Medical Center — UL1TR002014: Penn State Clinical and Translational Science Institute • The University of Michigan at Ann Arbor — UL1TR002240: Michigan Institute for Clinical and Health Research • Vanderbilt University Medical Center — UL1TR002243: Vanderbilt Institute for Clinical and Translational Research • University of Washington — UL1TR002319: Institute of Translational Health Sciences • Washington University in St. Louis UL1TR002345: Institute of Clinical and Translational Sciences • Oregon Health & Science University — UL1TR002369: Oregon Clinical and Translational Research Institute • University of Wisconsin-Madison — UL1TR002373: UW Institute for Clinical and Translational Research • Rush University Medical Center — UL1TR002389: The Institute for Translational Medicine (ITM) • The University of Chicago — UL1TR002389: The Institute for Translational Medicine (ITM) • University of North Carolina at Chapel Hill — UL1TR002489: North Carolina Translational and Clinical Science Institute • University of Minnesota — UL1TR002494: Clinical and Translational Science Institute • Children’s Hospital Colorado — UL1TR002535: Colorado Clinical and Translational Sciences Institute • The University of Iowa — UL1TR002537: Institute for Clinical and Translational Science • The University of Utah — UL1TR002538: Uhealth Center for Clinical and Translational Science • Tufts Medical Center — UL1TR002544: Tufts Clinical and Translational Science Institute • Duke University — UL1TR002553: Duke Clinical and Translational Science Institute • Virginia Commonwealth University — UL1TR002649: C. Kenneth and Dianne Wright Center for Clinical and Translational Research • The Ohio State University — UL1TR002733: Center for Clinical and Translational Science • The University of Miami Leonard M. Miller School of Medicine — UL1TR002736: University of Miami Clinical and Translational Science Institute • University of Virginia — UL1TR003015: iTHRIV Integrated Translational health Research Institute of Virginia • Carilion Clinic — UL1TR003015: iTHRIV Integrated Translational health Research Institute of Virginia • University of Alabama at Birmingham — UL1TR003096: Center for Clinical and Translational Science • Johns Hopkins University — UL1TR003098: Johns Hopkins Institute for Clinical and Translational Research • University of Arkansas for Medical Sciences — UL1TR003107: UAMS Translational Research Institute • Nemours — U54GM104941: Delaware CTR ACCEL Program • University Medical Center New Orleans — U54GM104940: Louisiana Clinical and Translational Science (LA CaTS) Center • University of Colorado Denver, Anschutz Medical Campus — UL1TR002535: Colorado Clinical and Translational Sciences Institute • Mayo Clinic Rochester — UL1TR002377: Mayo Clinic Center for Clinical and Translational Science (CCaTS) • Tulane University — UL1TR003096: Center for Clinical and Translational Science • Loyola University Medical Center — UL1TR002389: The Institute for Translational Medicine (ITM) • Advocate Health Care Network — UL1TR002389: The Institute for Translational Medicine (ITM) • OCHIN — INV-018455: Bill and Melinda Gates Foundation grant to Sage Bionetworks

### Additional Data Partners Who Have Signed a DTA and Whose Data Release is Pending

The Rockefeller University — UL1TR001866: Center for Clinical and Translational Science • The Scripps Research Institute — UL1TR002550: Scripps Research Translational Institute • University of Texas Health Science Center at San Antonio — UL1TR002645: Institute for Integration of Medicine and Science • The University of Texas Health Science Center at Houston — UL1TR003167: Center for Clinical and Translational Sciences (CCTS) • NorthShore University HealthSystem — UL1TR002389: The Institute for Translational Medicine (ITM) • Yale New Haven Hospital — UL1TR001863: Yale Center for Clinical Investigation • Emory University — UL1TR002378: Georgia Clinical and Translational Science Alliance • Weill Medical College of Cornell University — UL1TR002384: Weill Cornell Medicine Clinical and Translational Science Center • Montefiore Medical Center — UL1TR002556: Institute for Clinical and Translational Research at Einstein and Montefiore • Medical College of Wisconsin — UL1TR001436: Clinical and Translational Science Institute of Southeast Wisconsin • University of New Mexico Health Sciences Center — UL1TR001449: University of New Mexico Clinical and Translational Science Center • George Washington University — UL1TR001876: Clinical and Translational Science Institute at Children’s National (CTSA-CN) • Stanford University — UL1TR003142: Spectrum: The Stanford Center for Clinical and Translational Research and Education • Regenstrief Institute — UL1TR002529: Indiana Clinical and Translational Science Institute • Cincinnati Children’s Hospital Medical Center — UL1TR001425: Center for Clinical and Translational Science and Training • Boston University Medical Campus — UL1TR001430: Boston University Clinical and Translational Science Institute • The State University of New York at Buffalo — UL1TR001412: Clinical and Translational Science Institute • Aurora Health Care — UL1TR002373: Wisconsin Network For Health Research • Brown University — U54GM115677: Advance Clinical Translational Research (Advance-CTR) • Rutgers, The State University of New Jersey — UL1TR003017: New Jersey Alliance for Clinical and Translational Science • Loyola University Chicago — UL1TR002389: The Institute for Translational Medicine (ITM) • #N/A — UL1TR001445: Langone Health’s Clinical and Translational Science Institute • Children’s Hospital of Philadelphia — UL1TR001878: Institute for Translational Medicine and Therapeutics • University of Kansas Medical Center — UL1TR002366: Frontiers: University of Kansas Clinical and Translational Science Institute • Massachusetts General Brigham — UL1TR002541: Harvard Catalyst • Icahn School of Medicine at Mount Sinai — UL1TR001433: ConduITS Institute for Translational Sciences • Ochsner Medical Center — U54GM104940: Louisiana Clinical and Translational Science (LA CaTS) Center • HonorHealth — None (Voluntary) • University of California, Irvine — UL1TR001414: The UC Irvine Institute for Clinical and Translational Science (ICTS) • University of California, San Diego — UL1TR001442: Altman Clinical and Translational Research Institute • University of California, Davis — UL1TR001860: UCDavis Health Clinical and Translational Science Center • University of California, San Francisco — UL1TR001872: UCSF Clinical and Translational Science Institute • University of California, Los Angeles — UL1TR001881: UCLA Clinical Translational Science Institute • University of Vermont — U54GM115516: Northern New England Clinical & Translational Research (NNE-CTR) Network • Arkansas Children’s Hospital — UL1TR003107: UAMS Translational Research Institute

## AUTHOR INFORMATION

Authorship was determined using ICMJE recommendations.

Conceptualization - J.B., L.K., K.Y., J.T.J, S.P., J.M.F., R.J.A., J.O.B.; Data curation - J.B.; Formal Analysis - J.B.; Funding acquisition - J.B., R.J.A, J.O.B; Investigation - J.B., R.J.A; Methodology - J.B., J.M.F, R.J.A.; Project administration - J.B., R.J.A., J.O.B.; Resources - J.B., R.J.A; Software - J.B.; Supervision - R.J.A., J.O.B.; Validation - J.B.; Visualization - J.B.; Writing – original draft - J.B., J.M.F., R.J.A., J.O.B.; Writing – review & editing- J.B., L.K., K.Y., J.T.J., S.P., J.M.F., R.J.A., J.O.B.

## REFERENCES

1. Evans DG, Howard E, Giblin C, et al. Birth incidence and prevalence of tumor-prone syndromes: estimates from a UK family genetic register service. Am J Med Genet A. 2010;152A(2):327–332.

2. Dg E, Nl B, S T, et al. Schwannomatosis: a genetic and epidemiological study. J Neurol Neurosurg Psychiatry. 2018;89(11). doi:10.1136/JNNP-2018-318538

3. Uusitalo E, Rantanen M, Kallionpää RA, et al. Distinctive Cancer Associations in Patients With Neurofibromatosis Type 1. J Clin Oncol. 2016;34(17):1978–1986.

4. Rasmussen SA, Yang Q, Friedman JM. Mortality in neurofibromatosis 1: an analysis using U.S. death certificates. Am J Hum Genet. 2001;68(5):1110–1118.

5. Radtke HB, Klein-Tasman BP, Merker VL, et al. The impact of the COVID-19 pandemic on neurofibromatosis clinical care and research. Orphanet J Rare Dis. 2021;16(1):61.

6. Wolters PL, Reda S, Martin S, et al. Impact of the coronavirus pandemic on mental health and health care in adults with neurofibromatosis: Patient perspectives from an online survey. Am J Med Genet A. Published online September 18, 2021. doi:10.1002/ajmg.a.62490

7. Haendel MA, Chute CG, Bennett TD, et al. The National COVID Cohort Collaborative (N3C): Rationale, design, infrastructure, and deployment. J Am Med Inform Assoc. 2021;28(3):427–443.

8. Bennett TD, Moffitt RA, Hajagos JG, et al. Clinical Characterization and Prediction of Clinical Severity of SARS-CoV-2 Infection Among US Adults Using Data From the US National COVID Cohort Collaborative. JAMA Netw Open. 2021;4(7):e2116901.

9. Phenotype_Data_Acquisition Wiki. Github Accessed August 11, 2021. https://github.com/National-COVID-Cohort-Collaborative/Phenotype_Data_Acquisition

10. Incidence and Death Rates. Published June 8, 2021. Accessed July 27, 2021. https://www.cdc.gov/cancer/uscs/technical_notes/stat_methods/rates.htm

11. Anderson RN, Rosenberg HM. Age standardization of death rates: implementation of the year 2000 standard. Natl Vital Stat Rep. 1998;47(3):1–16, 20.

12. Efron B, Tibshirani RJ. An Introduction to the Bootstrap. CRC Press; 1994.

13. WHO Working Group on the Clinical Characterisation and Management of COVID-19 infection. A minimal common outcome measure set for COVID-19 clinical research. Lancet Infect Dis. 2020;20(8):e192–e197.

14. Gregory JM, Slaughter JC, Duffus SH, et al. COVID-19 Severity Is Tripled in the Diabetes Community: A Prospective Analysis of the Pandemic’s Impact in Type 1 and Type 2 Diabetes. Diabetes Care. 2021;44(2):526–532.

15. Lim S, Bae JH, Kwon HS, Nauck MA. COVID-19 and diabetes mellitus: from pathophysiology to clinical management. Nat Rev Endocrinol. 2021;17(1):11–30.

16. Brandt JS, Hill J, Reddy A, et al. Epidemiology of coronavirus disease 2019 in pregnancy: risk factors and associations with adverse maternal and neonatal outcomes. Am J Obstet Gynecol. 2021;224(4):389.e1-389.e9.

17. Richardson S, Hirsch JS, Narasimhan M, et al. Presenting Characteristics, Comorbidities, and Outcomes Among 5700 Patients Hospitalized With COVID-19 in the New York City Area. JAMA. 2020;323(20):2052–2059.

18. Barron E, Bakhai C, Kar P, et al. Associations of type 1 and type 2 diabetes with COVID-19-related mortality in England: a whole-population study. Lancet Diabetes Endocrinol. 2020;8(10):813–822.

19. Korf B. Considerations for Individuals with NF Regarding the Novel Coronavirus. Published March 26, 2020. Accessed August 12, 2021. https://www.uab.edu/medicine/nfprogram/blog/165-considerations-for-individuals-with-nf-regarding-the-novel-coronavirus

