## Supplementary Information for "COVID-19 in people with neurofibromatosis 1, neurofibromatosis 2, or schwannomatosis"

### SUPPLEMENTAL MATERIAL

#### Supplemental Methods

##### *Developing concept sets for people diagnosed with neurofibromatosis*

To identify people with NF1, we created concept sets broad enough to include various diagnosis codes associated with NF1, but specific enough to exclude patients who have diagnoses that may not be related to NF1. A diagnosis of NF1 is often correlated with heterogeneity in tumor types leading to several diagnosis codes in EHRs for a given patient. Heterogeneity in NF1 related codes can be especially problematic when patients are assigned diagnosis codes that relate to an older definition of the disease or associated tumor type. We closely evaluated the diagnosis codes being used by each site and defined a minimal set of NF1-related diagnosis codes to reduce the chance of including individuals without NF1 in the NF1 cohort. Supplemental Figure 1 shows the various terms (concept codes) that identify people with NF1 in our cohort and the variety of institutions where they were cared for. It shows that 86.3% of all patients selected using the NF1 concept set have a diagnosis of neurofibromatosis type 1 and 15.9% have the diagnosis of neurofibroma (Supplemental Figure 1). We then trimmed the concept set from 57 concepts (N3C concept set ID: 792972142) to 24 concepts (N3C concept set ID: 775352633) and found a difference of only 2 patients. We used the concept set with 24 terms to define our final cohort for analysis. Similar selection and trimming criteria were applied to concept sets for NF2 (N3C concept set ID: 880978583) and SWN (N3C concept set ID: 126122644) to generate the final concept sets (Supplemental Tables 2-3).

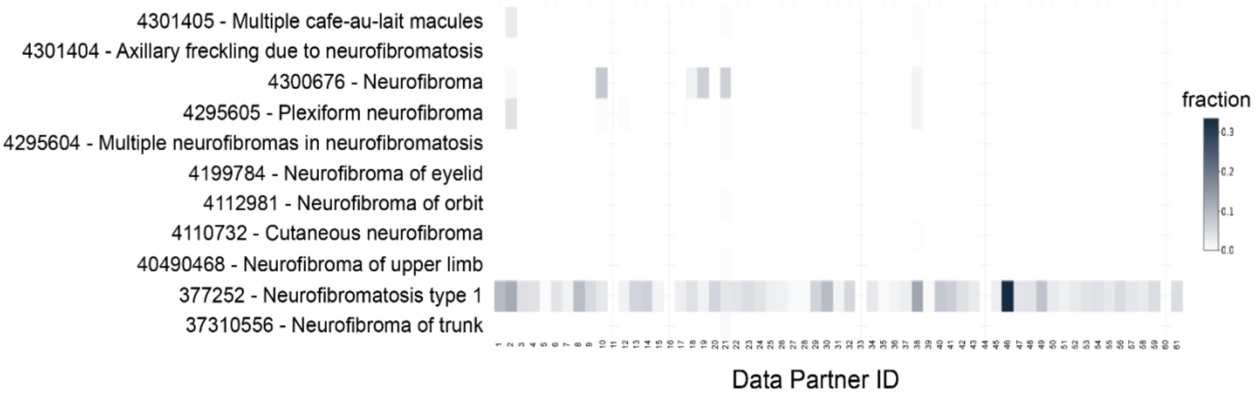


**Supplemental Figure 1: Concept Set of Neurofibromatosis Type 1.** A heatmap of the concept set used to generate the Neurofibromatosis Type 1 cohort is shown. The rows or y-axis consist of the various concept codes used to identify patients with Neurofibromatosis Type 1. The columns or x-axis represent various data partners who shared the patient EHR data. The dark colored boxes in the heatmap represent the number of patients from respective data partners in the x-axis that correspond to the concept code in the y-axis. The figure shows that most patients in the NF1 cohort correspond to concept codes of “Neurofibroma” and “Neurofibromatosis Type 1” (i.e., the majority of patients in the cohort are included by being associated with these two concept codes).

Supplemental Table 1 - NF1 Concept Set, the concept codes used to select the NF1 cohort in the N3C database.
Supplemental Table 2 - NF2 Concept Set, the concept codes used to select the NF2 cohort in the N3C database.
Supplemental Table 3 - SWN Concept Set, the concept codes used to select the SWN cohort in the N3C database.
Supplemental Table 4 - TS Concept Set, the concept codes used to select the TS cohort in the N3C database.
Supplemental Table 5 - FX Concept Set, the concept codes used to select the FX cohort in the N3C database.
Supplemental Table 6 - MCC Concept Set, the concept codes used to select the MCC cohort in the N3C database.
Supplemental Table 7 - AML Concept Set, the concept codes used to select the AML cohort in the N3C database.
Supplemental Table 8 - HYP Concept Set, the concept codes used to select the HYP cohort in the N3C database.
Supplemental Table 9 - DM1 Concept Set, the concept codes used to select the DM1 cohort in the N3C database.
Supplemental Table 10 - A demographic summary of the NF and non-NF cohorts.
Supplemental Table 11 – Age-adjusted counts of positive cases in different disease cohorts.
Supplemental Table 12 - Age-adjusted counts of severe outcomes in different disease cohorts.
Supplemental Table 13 - Age-adjusted counts of invasive ventilation in different disease cohorts.
Supplemental Table 14 - Median length of hospital stay in different disease cohorts for people with severe outcomes.
